# Sex and the clock: Exploring sex differences in chronotype and circadian preferences among cognitively healthy older adults

**DOI:** 10.1101/2024.11.26.24318021

**Authors:** Natalie S Pandher, Leslie Yack, Esther Li, Quentin Coppola, Kaitlin B Cassaletto, Lea T Grinberg, Thomas C Neylan, Joel H Kramer, Christine M Walsh

## Abstract

This study aimed to compare objective circadian rest-activity-rhythm (RAR) measures with self-reported circadian behavior and morning-evening preference in cognitively healthy older men and women. A total of 129 participants (ages 65-90) completed the Horne & Ostberg Morning-Eveningness Questionnaire (MEQ) and the Circadian Type Inventory (CTI) to assess their morning-evening preference and circadian traits, including rigidity, vigor, languidness, and flexibility. These subjective measures were compared to objective actigraphy data from a sub-cohort of 70 individuals who wore actigraphy watches for 24 hours a day over a 7-day period. Rest-activity rhythm variables were derived from the actigraphy data and analyzed. The results revealed sex differences in circadian behavior. Languid and flexible men (higher CTI scores) had significantly higher depression scores (GDS, p = 0.045) and showed more evening preference (MEQ, p = 0.007) compared to their vigorous and rigid counterparts, while no significant differences were found in women. Actigraphy data showed that men had less stable (interdaily stability, IS, p = 0.030) and more fragmented (interdaily variability, IV, p = 0.001) circadian rhythms than women. A significant association was found between acrophase time and CTI-FR scores (β = −0.406, p < 0.001), with earlier acrophase times linked to more rigid circadian rhythms. Additionally, earlier acrophase times and morningness preference were associated with faster information processing speeds (IPS, p = 0.018). These findings suggest that circadian behaviors and their impact on mood and cognition are more pronounced in older men than women, highlighting the value of actigraphy in assessing circadian profiles and sex differences in aging.

## Introduction

Maintaining a healthy circadian rhythm is critical for optimal sleep, mood, cognitive function, and overall health. Circadian rhythms are endogenous, 24-hour cycles governing many physiological processes, and the disruption of these rhythms can lead to various homeostatic imbalances (Lack & Wright, 2007). Existing literature has established that circadian rhythms become dampened and advanced with old age, as older adults exhibit lower amplitudes, earlier acrophase times, and less day to day variability when compared with younger populations (Erickson et al., 2024; Li et al., 2018; J. A. Mitchell et al., 2017). These rhythm changes have been shown to contribute to increased odds of developing dementia and mild cognitive impairment (MCI) in older adults collectively, as well as in older women and men separately (Musiek et al., 2018; Rogers-Soeder et al., 2018; Tranah et al., 2011). Much of the literature thus far has focused on objectively measured circadian behavior by methods such as wrist-based actigraphy, core body temperature monitoring, or hormone fluctuation. Subjective measures of circadian patterns such as morning vs evening preference or sleep schedule patterns have not been studied extensively in the older adult population. It is also unclear how perceived circadian patterns, and objectively measured ones, might relate to each other. Understanding the interplay between subjective and objective measures of circadian rhythm in an aging but cognitively healthy population can provide valuable insights into associations of cognitive health, and potential disease modifiers.

The Morningness-Eveningness Questionnaire (MEQ) is one well-established, subjective, chronotype questionnaire that categorizes an individual into a morning type, evening type, or neither type person based on their time of day preference (Horne & Ostberg, 1976). Previous research on older adults has explored relationships between age and time of day preference using the MEQ. A study focusing on age related differences between a cohort of healthy older adults (mean age of 83 years) matched to an equal number of young controls (mean age of 25) found that older adults scored higher on the MEQ (indicating stronger morning preference) than their younger counterparts (Monk et al., 1991). This finding by Monk et al. (1991) is supported by the results of a later study done by Carrier et al. (1997) in “middle age” adults (20-59 years of age) in which aging was associated with an increase in morningness. None of the literature thus far has found a significant gender effect on aging and chronotype.

Another validated, circadian questionnaire administered widely is the Circadian Type Inventory (CTI), which assesses an individual’s daytime energy levels on a scale of languidness to vigor and sleep schedule flexibility on a scale of flexible to rigid (Di Milia et al., 2004; Smith et al., 1993). Research using the CTI has been performed predominantly in shift-working populations as the CTI is often used to assess an individual’s propensity for handling shift work or changing their sleep schedule (Folkard et al., 1979). However, there have been a few landmark studies looking at aging and the constructs of circadian vigor or rigidity. Monk et al. (1991) used the Circadian Type Questionnaire (Folkard et al., 1979), an older version of the current CTI to highlight a link between aging and greater daytime vigor and sleep schedule rigidity (Monk et al., 1991). Marcoen et al. (2015) performed a study on participants ages 18 to 83 (N= 752) and found a significant gender effect in CTI scores with men demonstrating greater sleep schedule flexibility than women (Marcoen et al., 2015). They also found a negative correlation between subjective circadian flexibility and morningness, indicating that people with more flexible sleep schedules tend towards evening preference while those with more rigid schedules tend toward morningness (Marcoen et al., 2015). However, morningness here was assessed by the CSM or Composite Scale of Morningness (Folkard et al., 1979; Torsvall & Åkerstedt, 1980), not the MEQ (Horne & Ostberg, 1976).

Of these studies using the MEQ and CTI, only a handful include objectively measured circadian data (Carrier et al., 1997; Monk et al., 1991), and neither of these explore wrist-based actigraphy to assess circadian rhythm, instead using invasive in-lab PSG protocols. The existing literature on wrist-based actigraphy and circadian patterns in older adults shows a dampening of overall rhythm with less daytime activity and decreased robustness (de Feijter et al., 2020; Erickson et al., 2024; Li et al., 2021). Studies in older adults highlight earlier acrophase times, or peak activity occurring earlier in the day, more stable, and less fragmented rhythms (Li et al., 2021; J. A. Mitchell et al., 2017). The current literature looking specifically at older men and women is focused on relationships between RAR and risk of cognitive decline or dementia. In an actigraphy-based study, older adult women with delayed acrophase times (after 3:51 PM) had a greater risk for developing cognitive impairment or displaying signs of poor cognition (Tranah et al., 2011; Walsh et al., 2014). Conversely, older adult men with advanced acrophase (time of peak activity occurring before 12:28pm) had a higher risk of experiencing significant cognitive decline (Rogers-Soeder et al., 2018). This finding seems to contradict prior research showing a link between delayed acrophase and symptoms of cognitive decline in older adults (Cochrane et al., 2012). The contradictory nature of the current body of research on objective circadian rhythms in older adults highlights its complexity and emphasizes the importance of considering both subjective and objective measures, along with age and gender differences, to understand individual variations in circadian rhythms.

Much of the existing research on the MEQ and CTI has focused on younger populations and shift workers, and actigraphy has not been used as an objective measure alongside these subjective circadian questionnaires in older adult populations as of yet. This paper investigates the relationship between subjective and objective assessments of circadian rhythmicity in a sample of healthy older adults by focusing on key activity patterns that may influence cognitive function. We utilized wrist-based actigraphy, a non-invasive method that objectively measures circadian rhythmicity, and we also looked at the relationship between these circadian measures and cognitive processing speed (IPS). IPS has previously been shown to slow with age and is associated with white matter integrity (Kerchner et al., 2012; Sweet, 2011). Lastly, we looked closely at gender differences in these variables and how older men and women vary in their reports of circadian preference and type. Our study addresses the key question: Does combining subjective and objective data provide a better understanding of circadian rhythm in cognitively healthy older men and women? By exploring this question, we aim to contribute to a deeper understanding of how circadian rhythms influence cognitive health and overall well-being in older adults. Understanding how circadian patterns are associated with sex, age, and cognition is essential for developing modifiable interventions to maintain or enhance cognitive health in the aging population.

## Methods

### Participants

129 cognitively healthy, community-dwelling, older adults between the ages of 65 and 90 years were enrolled from the Brain Aging Network for Cognitive Health (BrANCH) study at the University of California, San Francisco (UCSF) Memory and Aging Center (enrollment year range: 2012-2023). These participants were originally recruited into BRANCH via flyers, newspaper advertisements, and community outreach events. Through the BrANCH study, all participants completed a general research visit which included neuropsychological testing, neurological examination, and a Clinical Dementia Rating (CDR) completed via interview with a study partner or informant. A “cognitively normal” consensus-based diagnosis was assigned by a formal committee if at least two of the following criteria were met: no cognitive concerns during the neurological examination, a participant performed within expected ranges for their age and education on neuropsychological assessments, or the study informant or partner did not raise any concerns over the participant’s cognition during the interview. Participants were excluded if they had severe psychiatric illness, neurologic disorders (e.g., epilepsy, multiple sclerosis), or medical conditions that could impact cognition (e.g., recent substance use disorders, active chemotherapy). Participants that were classified as cognitively normal were then referred to the sleep study. If participants agreed to participate, they and their partners/informants, were asked to provide written, informed consent. Study protocols and consent forms were approved by the UCSF Committee on Human Research (IRB #11-07991).

### Circadian questionnaires

To assess morning or evening preference, participants completed the Horne & Ostberg Morningness-Eveningness Questionnaire (Horne & Ostberg, 1976). The Morningness-Eveningness Questionnaire (MEQ) is a 19-item questionnaire that identifies where along a morning-type to evening-type scale an individual is, based on the times of day during which the participant reports feeling most active and alert. The questions on the MEQ are a combination of Likert-type items and simple time scales from which participants can choose the ideal span of time during which they would complete various tasks requiring either intense focus, physical labor, or nighttime work. In their first validation of the MEQ, Horne and Ostberg (1979) compressed MEQ total scores into a five-point morningness-eveningness scale using these categories: definitely morning type (70–86), moderately morning type (59–69), neither type (42– 58), moderately evening type (31–41), and definitely evening type (16–30). However, this categorization did not accurately represent our sample of predominantly morning type individuals with very few definitely evening types. To address this issue, we collapsed across moderate and definite evening type groups to form one “evening type”. Thus, analyses were performed for four groups: definitely morning type (70-86), moderately morning type (59–69), neither type (42-58), and evening type (16-41). The MEQ has been used previously in older adult populations, but our aim in utilizing it in this study was to correlate it with the CTI and actigraphy measures which has not been done before.

Participants also completed the 11-item Circadian Type Inventory (CTI), which is a Likert-based questionnaire that assesses an individual’s adaptability to sleep pattern changes by looking at rhythm stability and amplitude (Di Milia et al., 2004; Smith et al., 1993; Folkard & Monk, 1979). Rhythm stability is measured on the FR scale where an individual indicates how flexible or rigid they are when faced with sleep schedule changes (Di Milia et al., 2004). Rhythm amplitude is reflected by the LV scale where an individual reports how languid or vigorous they are in the daytime, especially when faced with reduced sleep (Di Milia et al., 2004). Lower scores on the FR scale are representative of circadian rigidity or a decreased ability to deviate from a set sleep schedule, and higher scores indicate flexibility, meaning an improved ability to deviate from a set schedule (Di Milia et al., 2004). Similarly, low LV scores exhibit vigor or higher daytime energy even after a poor night of sleep and high LV scores are associated with languidness or lethargy following a night of reduced sleep (Di Milia et al., 2004; Smith et al., 1993). Our goal in utilizing the CTI was to determine subjective understanding of circadian traits in an aging population by asking for self-reported behaviors that characterize an individual’s sense of rhythm stability (FR) and amplitude (LV).

### Cognitive function measurements

In a sub cohort of eighty-five participants, a battery of cognitive tests assessing information processing speed (Kerchner et al., 2012) were administered within 90 days of the circadian questionnaires and actigraphy measurements. The reaction time in seconds, or response latency, for each of the seven visuospatial tasks that made up the testing battery was combined to create a composite score, and these composite scores were used in our analysis to reflect information processing speed (IPS) (Kerchner et al., 2012; Sweet, 2011).

In addition to IPS, we also used Mini-Mental State Examination (MMSE) scores as a marker of cognitive function in our sample (A. J. Mitchell, 2009). The MMSE consists of 19 tests across 11 domains, including orientation, registration, attention, calculation, recall, naming, repetition, comprehension, writing, and construction (Folstein et al., 1975). Scores ranging from 25-30 are indicative of normal cognitive status, and we used this as an exclusion criteria to ensure that all participants had baseline normal cognitive function (MMSE>27). Additionally, including MMSE scores as a covariate in our analyses served to remove uncertainty that individuals with different MMSE scores had lower levels of cognitive function, less presence of mind, intellectual disability, or mental confusion during the IPS battery or while answering questionnaires.

### Seven-day actigraphy assessment

Participants were asked to wear a wireless actigraphy watch on either their dominant or nondominant wrist for seven days and nights. Recording time each day was up to 24 hours as participants were told to only remove the watch for water-related activities such as bathing and swimming. 35 participants who took part in the study from 2012 to 2019 wore the Micro Motionlogger Sleep Watch (Ambulatory Monitoring Inc, NY), while the 38 participants seen from 2019 to 2023 wore the Philips Respironics Actiwatch Spectrum Plus (Phillips-Respironics, Bend, Oregon). An actigraphy recording was considered valid if it had no more than four hours of lost data in any twenty-four-hour period, and if there were at least three valid twenty-four-hour periods in the data set. These criteria followed those previously published in older adults (Ancoli-Israel et al., 2003; Blackwell et al., 2017; Walsh et al., 2014). Data acquired using the AMI Micro Motionlogger were processed using ActionW4 (Ambulatory Monitoring Inc, NY) to garner nonparametric rest-activity-rhythm variables. Data acquired using the Actiwatch Spectrum Plus were analyzed using Philips Actiware (Phillips-Respironics, Bend, Oregon) and R packages “mice” (Buuren et al., 2023), “RAR” (Graves, 2019/2023), and “nparACT” (Schabus, 2017). These scripts were used to derive the rest-activity-rhythm (RAR) variables used in the analysis, and the definitions and derivation of the RAR variables of interest (Cavalcanti-Ferreira et al., 2018; de Feijter et al., 2020; Feng et al., 2023; Gonçalves et al., 2014) are presented in the table below.

Rest-activity rhythm variables derived by nonparametric analysis.

Rest-activity rhythm variables derived by nonparametric analysis.
| RAR Variable | Definition | Derivation | Interpretation |
| --- | --- | --- | --- |
| Interdaily Stability (IS) | a measure of the strength of coupling of a rhythm to environmental zeitgebers | ratio between the variance of the average 24-hour pattern around the mean and the overall variance | High IS = stable rhythm<br>Low IS = less stable rhythm |
| Intradaily Variability (IV) | a measure of fragmentation of the circadian rhythm | ratio of the mean squares of the difference between successive hours and the mean squares around the grand mean | High IV = fragmented rhythm<br>Low IV = intact rhythm |
| Relative Amplitude (RA) | the difference between activity levels during the most and least active times of day | average intensity of the most active 10 hours (*M10*) minus the average intensity of the least active 5 hours (*L5*) divided by $M10 + L5$ | High RA = pronounced rest-activity rhythm<br>Low RA = less pronounced rhythm |
| Acrophase | time of peak activity occurring during the 24-hour day | time at which cosinor curve fitted to the circadian rhythm reaches its peak | Earlier than 1:30pm = *advanced phase*<br>1:30pm to 3:50pm = *normal phase*<br>After 3:50pm = *delayed phase* |

### Statistical Analysis

Data were analyzed using IBM SPSS Statistics Version 29.01.00 (171). Data points three standard deviations from the mean were considered outliers and removed from the dataset. Additionally, for the rest-activity-rhythm variables (IV, IS, RA, acrophase) we assessed distribution across the two watches to ensure variables were not affected by watch type. The study design resulted in some missing data from participants. To maximize the data used, participants were only excluded from analyses where the missing data directly impacted the results. They were included in all other analyses. Our variables of interest had kurtosis and skewness values close to zero (within a −2 to +2 range) prior to normalization, so we opted for analysis on the dataset without normalizing. Our hypothesis involved finding direct relationships between subjective (circadian questionnaires) and objective (actigraphy) measures, so we used a multiple linear regression approach covarying for age, gender, and MMSE. To assess age and gender effects, we used one-way ANOVAs, independent sample T-tests, and linear regressions (when age was a continuous variable). Instead of including medication use as a separate covariate and reducing statistical power, we created a third gender category for the participants (all female) that were taking sleep modifying medications and included medication use as a covariate in all analyses including female participants. When utilizing the actigraphy data, we covaried for the type of watch worn by the participant, AMI or Philips, to reduce any device-dependent data bias. As a final check, dependent variables were plotted against residuals, and a Q-Q plot was generated for each to ensure variables of interest had normal distributions. The Durbin Watson test (d_u_ < d< 4-d_u_) was also performed on significant relationships to rule out autocorrelation as a possible reason for the trends observed. Statistically significant relationships were defined as correlations with a p-value ≤ 0.05.

## Results

### Demographics

Participants had a mean age of 74.66 ± 5.35 years and mean MMSE scores of 29.03 ± 1 (Table 1). A total of five participants, all females, took sleep-modifying medications during the study period. These were antidepressants of the following drug classes: tri-cyclic (TCA), selective serotonin reuptake inhibitor (SSRI), and selective serotonin and norepinephrine reuptake inhibitor (SNRI).

**Table 1.**
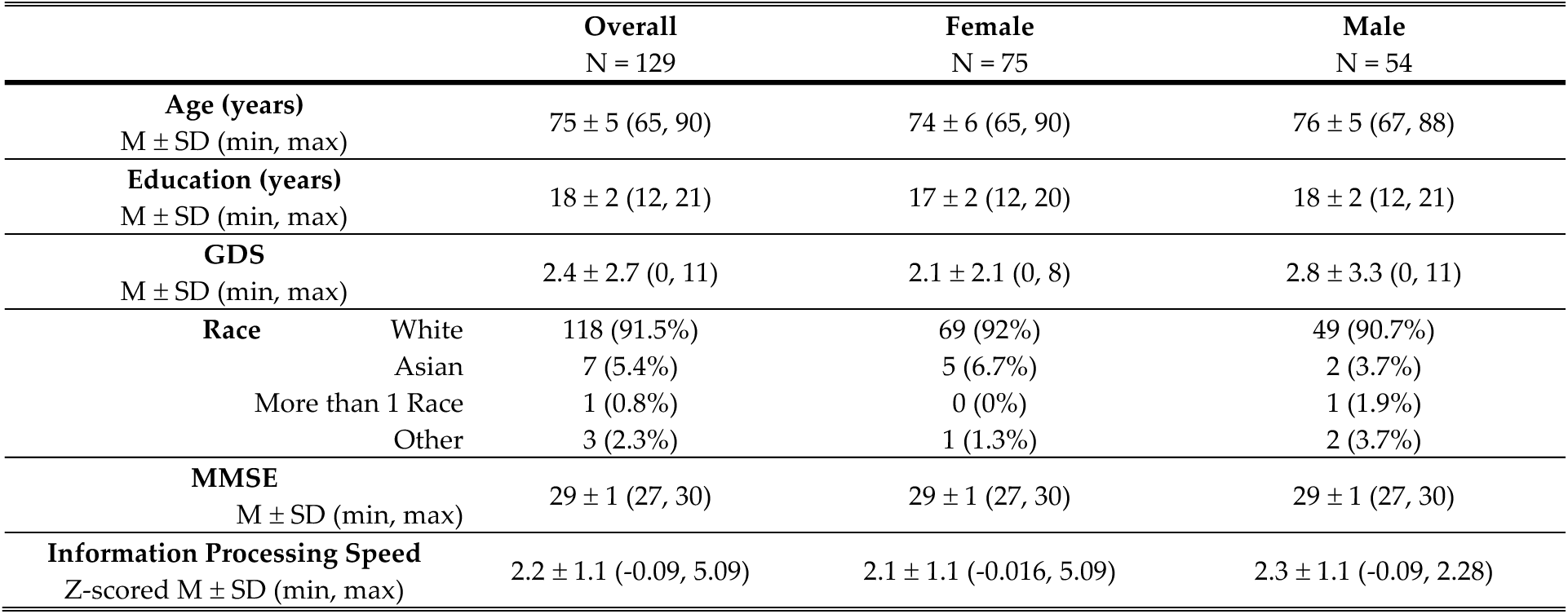
Demographics, mood, and cognitive function scores in males, females, and across the overall sample.

### Gender Differences in Subjective Circadian Measures

#### Circadian Type Inventory

Across the entire sample, participants had an average CTI-LV score of 13.52 ± 4.16 and CTI-FR score of 12.6 ± 4.15. LV scores range from 6 at the lowest and 30 at the highest, with higher scores indicating more languidity. We used the midpoint of 6 and 30, as the cutoff to separate our sample into two groups: those identifying as languid individuals (LV=19-30) and those as vigorous individuals (LV=6-18). The FR score range is between 5 to 25 with higher scores indicating more flexibility, so again using the midpoint, we determined the flexible (FR=16-25) and rigid groups (FR=5-15). The healthy older adults in our sample had low LV and FR scores showing a propensity towards rigid sleeping schedules (n=93) and vigorous dispositions (n=107), rather than flexible schedules (n=29) and languid behaviors (n=15). We did not see any significant mean differences when comparing between the languid versus vigorous and the flexible versus rigid types due to the skewed sample. There was also no association found between age and CTI scores or gender and CTI scores. However, there were within-gender differences.

A within gender t-test between the circadian types showed that languid males had significantly higher scores on the Global Depression Scale (*p*=.045), lower scores on the Mini-Mental Status Examination (*p=*.013), and lower scores on the Morningness Eveningness Questionnaire (*p*=.007) than vigorous males. In women, there were no significant differences between the languid and vigorous groups. Similarly, men with flexible sleep schedules had MMSE (*p=*.001) and MEQ scores (*p*=.05) that were significantly different from men with rigid sleep schedules, while there were no mean differences between flexible and rigid women. These results suggest that circadian type may influence older men more than older women, and languid and flexible men may be more susceptible to low mood, eveningness, and cognitive issues than languid and flexible women. The within-gender comparisons for both LV and FR scores are summarized in Table 2.

**Table 2.** Characteristics of languid-vigorous and flexible-rigid types compared within gender. 2a. Within gender differences in languid and vigorous circadian group types. 2b. Within gender differences in flexible and rigid circadian group types.

| CTI-LV |  |  |  |  |  |  |  |  |  |  |
| --- | --- | --- | --- | --- | --- | --- | --- | --- | --- | --- |
|  | Male |  |  |  |  |  | Female |  |  |  |
|  | Languid<br>(n=8) | Vigorous<br>(n=43) | T-test |  |  | Languid<br>(n=7) | Vigorous<br>(n=64) | T-test |  |  |
|  |  |  | t | p-value | Mean Diff |  |  | t | p-value | Mean Diff |
| Age (years) | M = 76.4<br>SD = 3.26 | M = 75.4<br>SD = 4.90 | 0.571 | .520 | 1.03 | M = 73.8<br>SD = 6.64 | M = 73.9<br>SD = 5.79 | -0.038 | .970 | -0.088 |
| GDS | M = 5.00<br>SD = 5.13 | M = 2.29<br>SD = 2.53 | 1.36 | .045* | 2.17 | M = 3<br>SD = 2.94 | M = 2<br>SD = 2.1 | 0.892 | .376 | 1 |
| MMSE | M = 28.0<br>SD = 0.816 | M = 29.2<br>SD = 1.07 | -2.65 | .013* | -1.16 | M = 30<br>SD = 0 | M = 29.18<br>SD = 0.942 | 1.217 | .231 | 0.821 |
| MEQ | M = 54.3<br>SD = 11.4 | M = 64.5<br>SD = 7.73 | -2.84 | .007** | -10.15 | M = 55.17<br>SD = 8.78 | M = 61.92<br>SD = 7.99 | -1.959 | .054 | -6.753 |
| Information<br>Processing Speed | M = 2.83<br>SD = 1.49 | M = 2.07<br>SD = 0.879 | 1.88 | .068 | 0.758 | M = 1.39<br>SD = 0.587 | M = 2.15<br>SD = 1.16 | -0.916 | .365 | -0.759 |
| Acrophase Time | M = 15:03<br>SD = 1:12 | M = 14:22<br>SD = 1:10 | 1.151 | 0.130 | 0:40 | M = 14:25<br>SD = 1:41 | M = 14:20<br>SD = 0:57 | 0.164 | .435 | 0:05 |

2b. Within gender differences in flexible and rigid circadian group types.
| CTI-FR |  |  |  |  |  |  |  |  |  |  |
| --- | --- | --- | --- | --- | --- | --- | --- | --- | --- | --- |
| Male |  |  |  |  |  | Female |  |  |  |  |
|  | Flexible<br>(n=13) | Rigid<br>(n=38) | T-test |  |  | Flexible<br>(n=16) | Rigid<br>(n=55) | T-test |  |  |
|  |  |  | t | p-value | Mean Diff |  |  | t | p-value | Mean Diff |
| Age (years) | M = 75.6<br>SD = 4.85 | M= 75.5<br>SD = 4.67 | 0.062 | .951 | 0.095 | M = 74.7<br>SD = 6.46 | M = 73.65<br>SD = 5.68 | 0.656 | .514 | 1.092 |
| GDS | M = 3.38<br>SD = 3.85 | M = 2.63<br>SD = 3.14 | .567 | .574 | 0.742 | M = 1.64<br>SD = 1.5 | M = 2.19<br>SD = 2.29 | -0.752 | .455 | -0.550 |
| MMSE | M = 29.7<br>SD = 0.500 | M = 28.61<br>SD = 1.16 | 3.607 | .001** | 1.058 | M = 29.5<br>SD = 0.76 | M = 29.15<br>SD = 0.972 | 0.944 | .351 | 0.369 |
| MEQ | M = 57.7<br>SD = 12.1 | M = 65.3<br>SD = 6.31 | -2.162 | .048* | -7.599 | M = 58.3<br>SD = 6.57 | M = 62.11<br>SD = 8.48 | -1.82 | .080 | -3.825 |
| Information Processing Speed | M = 2.52<br>SD = 1.34 | M = 2.11<br>SD = 0.928 | 1.080 | .287 | 0.402 | M = 1.96<br>SD = 0.64 | M = 2.15<br>SD = 1.24 | -0.468 | .642 | -0.2 |
| Acrophase Time | M = 15:02<br>SD = 1:04 | M = 14:15<br>SD = 1:11 | 1.686 | 0.052 | 0:46 | M = 14:39<br>SD = 1:25 | M = 14:17<br>SD = 0:57 | 0.829 | .206 | 0:22 |

\*p<.05, \*\*p<.005

#### Morningness-Eveningness Questionnaire

The average MEQ score in this sample was 61.55 ± 9.53 with the caveat that most participants reported morning preference (n=85) or no preference (n=31) and only a handful reported evening preference (n=5). This propensity is supported by previous findings of individuals becoming more morning-type as they age (Carrier et al., 1997; Monk et al., 1991). We did find a weakly significant negative correlation (*p=*.045) between morning-evening preference and age in this older population with older adults tending towards evening preferences while the younger subset of the sample (65-75 years) reported morning preferences.

There was no association between gender and MEQ scores, but a within-gender ANOVA showed that in males, GDS scores, information processing speed, and acrophase time were significantly different between the chronotype groups. There was only one male participant that reported evening preference, so this data point was excluded from the ANOVA. Moderately morning-type men had higher depression scores (*p*=.022) and a delayed acrophase time (*p*=.004) when compared with definitely morning-type men. However, moderately morning-type men had faster information processing speeds than the other two groups (*p*=.017). This could suggest that morning preference is associated with better mood and peak activity earlier in the day, but less so with faster cognitive speeds. In females, there were no significant differences across the various preference groups. ANOVA results are summarized by gender and chronotype in Table 3.

**Table 3.** One-way ANOVA of morning vs evening types in males and females.

| Morningness-Eveningness Questionnaire |  |  |  |  |  |  |  |
| --- | --- | --- | --- | --- | --- | --- | --- |
| Male |  |  |  |  |  |  |  |
|  | Definitely<br>Morning-Type<br>n = 8 | Moderately<br>Morning-Type<br>n = 30 | Neither<br>Type<br>n = 11 | Evening<br>Type<sup>1</sup><br>n = 1 | F | p-value | η<sup>2</sup> |
| Age (years) | 75.4 ± 5.02 | 75.24 ± 4.62 | 76.01 ± 4.93 | 79.75 | 0.341 | 0.796 | 0.22 |
| GDS | 1.57 ± 1.72 | 2.86 ± 3.21 | 1.43 ± 2.29 | 11 | 3.683 | 0.022* | 0.251 |
| MMSE | 28.8 ± 1.09 | 29.1 ± 1.13 | 29 ± 1.27 | 29 | 0.064 | 0.979 | 0.007 |
| Info Processing Speed | 2.36 ± 1.35 | 1.97 ± 0.79 | 2.51 ± 0.82 | 5.07 | 3.894 | 0.017* | 0.25 |
| Acrophase Time | 13:22 ± 1:24 | 14:31 ± 0:39 | 15:29 ± 1:17 | 16:28 | 5.826 | 0.004** | 0.411 |
| Female |  |  |  |  |  |  |  |
|  | Definitely<br>Morning-Type<br>n = 11 | Moderately<br>Morning-Type<br>n = 36 | Neither<br>Type<br>n = 20 | Evening<br>Type<br>n = 4 | F | p-value | η<sup>2</sup> |
| Age (years) | 72.2 ± 5.227 | 74.28 ± 5.78 | 73.34 ± 5.264 | 81.25 ± 7.026 | 2.707 | 0.052 | 0.108 |
| GDS | 1.44 ± 1.13 | 2 ± 2.094 | 3 ± 2.66 | 1.67 ± 1.528 | 1.176 | 0.329 | 0.067 |
| MMSE | 29.25 ± 0.707 | 29.06 ± 0.998 | 29.25 ± 1.055 | 28.5 ± 2.121 | 0.381 | 0.767 | 0.031 |
| Info Processing Speed | 1.92 ± 0.950 | 2.07 ± 0.959 | 2.33 ± 1.48 | 3.04 ± 2.92 | 0.609 | 0.613 | 0.042 |
| Acrophase Time | 14:20 ± 0:47 | 14:11 ± 0:49 | 14:35 ± 1:25 | 15:33 ± 0:20 | 1.691 | 0.186 | 0.121 |

\*p<.05, \*\*p<.005
<sup>1</sup>Single data point for male evening-type was not included in ANOVA.

### Comparing across subjective circadian type and morning preference

To investigate how circadian type and morning-evening preference are related, we compared CTI scores and MEQ total scores using a multiple linear regression model covarying for age, gender, and medication use (Table 4). Both the CTI-LV (β=-0.406, *p*<.001) and CTI-FR (β=-0.228, *p*=.013) were significantly associated with MEQ scores, indicating that morning-type individuals tended to be rigid and vigorous circadian types. In women, low CTI-LV scores (β=-0.239, *p*=.041) were weakly associated with morningness on the MEQ (Figure 1a). In men, both scales of the CTI (LV: β=-0.612, *p*<.001, FR: β=-0.349, *p*=.015) were strongly predictive of time-of-day preference (Figure 1b). These findings illustrate that men with a morning preference are more likely to feel vigorous or experience more energy/wakefulness in the daytime and have a rigid circadian schedule that is resistant to change, while evening-type men experience the opposite with languid feelings in the day and more variable circadian schedules. Interestingly, this finding may not be as applicable for cognitively healthy older women suggesting that circadian-type behaviors are less dependent on chronotype.

**Figure 1.**
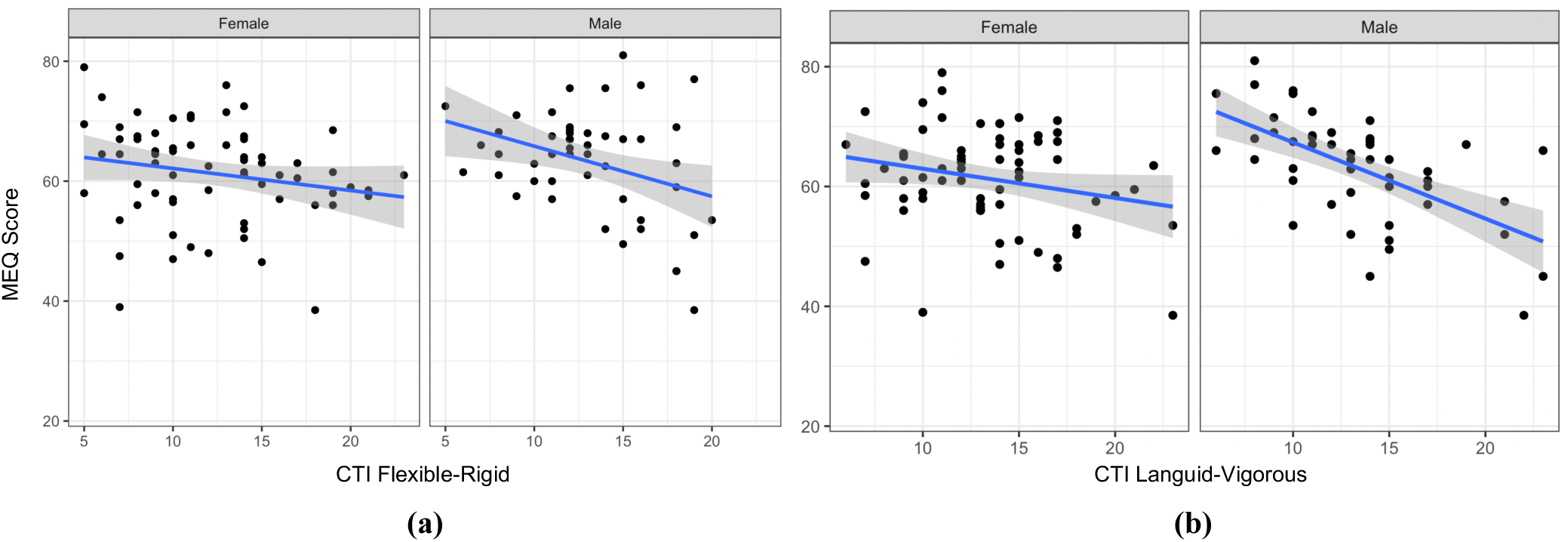
Associations between circadian type and morningness-eveningness vary across males and females. The relationship between **(a)** circadian rigidity and morning preference and **(b)** circadian vigor and morning preference is more significant in cognitively healthy older males (CTI-FR: R^2^ =.127; CTI-LV: R^2^ =.378) than in older females (CTI-FR: R^2^ =.119, CTI-LV: R^2^ =.159).

**Table 4.** Linear regression models of CTI and MEQ scores across the overall sample and within gender.

| Dependent Variable: Morningness-Eveningness Score (MEQ) |  |  |  |  |  |
| --- | --- | --- | --- | --- | --- |
| | | $\beta$ | t | p-value | Co-variates |
| **Overall**<br>N=117 | CTI-LV | -0.406 | -4.891 | <.001** | 1. Age, 2. Gender,<br>3. Medication Use |
|  | CTI-FR | -0.228 | -2.519 | 0.013* |  |
| **Female**<br>N=68 | CTI-LV | -0.239 | -2.084 | 0.041* | 1. Age, 2. Medication Use<sup>2</sup> |
|  | CTI-FR | -0.134 | -1.105 | 0.273 |  |
| **Male**<br>N=49 | CTI-LV | -0.612 | -5.255 | <.001** | 1. Age |
|  | CTI-FR | -0.349 | -2.535 | 0.015* |  |

<sup>1</sup>Sleep modifying medications (SSRIs, SNRIs, and TCAs) happened to be taken only by women in this sample.
\* $p<.05$
\*\* $p<.005$

### Objective Measures of Circadian Rhythmicity

Out of the 129 total participants, 73 individuals recorded actigraphy data by wearing either the Philips Spectrum Plus Actiwatch or the AMI Micro Motionlogger Sleep Watch. Demographics and rest-activity-rhythm (RAR) variables for all participants with usable actigraphy data are summarized in Table 5. The main variable of interest in our circadian analysis was acrophase: the time of day representing an individual’s peak level of activity and alertness. We computed acrophase in 24-hour clock time by converting radians to time and plotted the spread of acrophase time across our sample. Average acrophase time was 14:30:58 ± 00:01:03, with the earliest peak being 11:51:43 (11:52am) and the latest being 17:12:28 (5:12pm). To classify individuals into phase categories, we used acrophase cutoffs previously reported in older adults (Rogers-Soeder et al., 2018; Tranah et al., 2011), where an acrophase time earlier than 13:30 (1:30pm) is advanced phase, an acrophase later than 15:50 (3:50pm) constitutes a delayed phase, and an acrophase falling between 13:30 to 15:50 is a normal phase. We found that a majority of our sample (32 women and 23 men) fell in the normal phase category. This could be indicative of older adults generally having more time to engage in physical activity in the midafternoon, or it could be that these individuals preferred to be active in the sunniest or warmest part of the day.

**Table 5.**
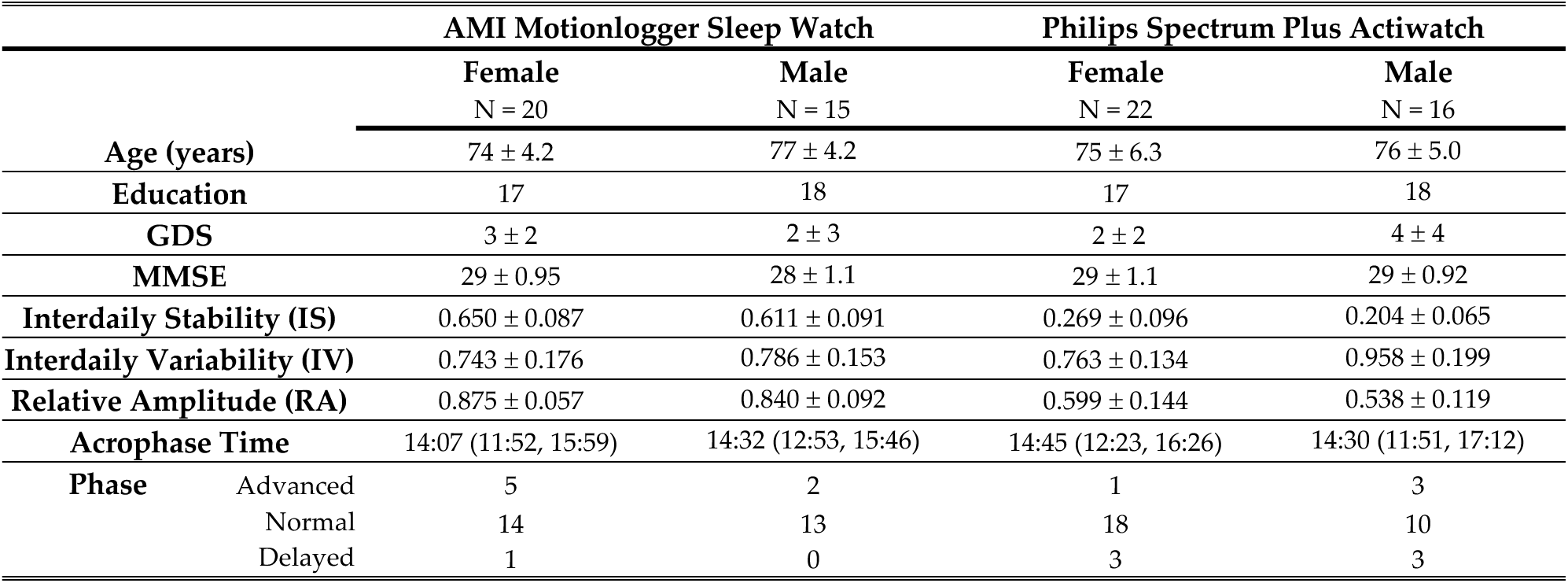
Actigraphy summary variables and demographic information displayed by watch type and gender.

#### Age and Rest Activity Rhythms

After combining the RAR variables that were comparable across watches (interdaily variability and acrophase time), we found a weakly significant correlation between age and acrophase time. Using a multiple linear regression model covarying for gender, medication use, and watch type, we found that age could be a significant predictor for acrophase time (β=.290, *p*=.016). Acrophase became more delayed in older individuals (Figure 2), supporting the shift towards lack of preference/evening preference in old age seen in MEQ score trends as well.

**Figure 2.**
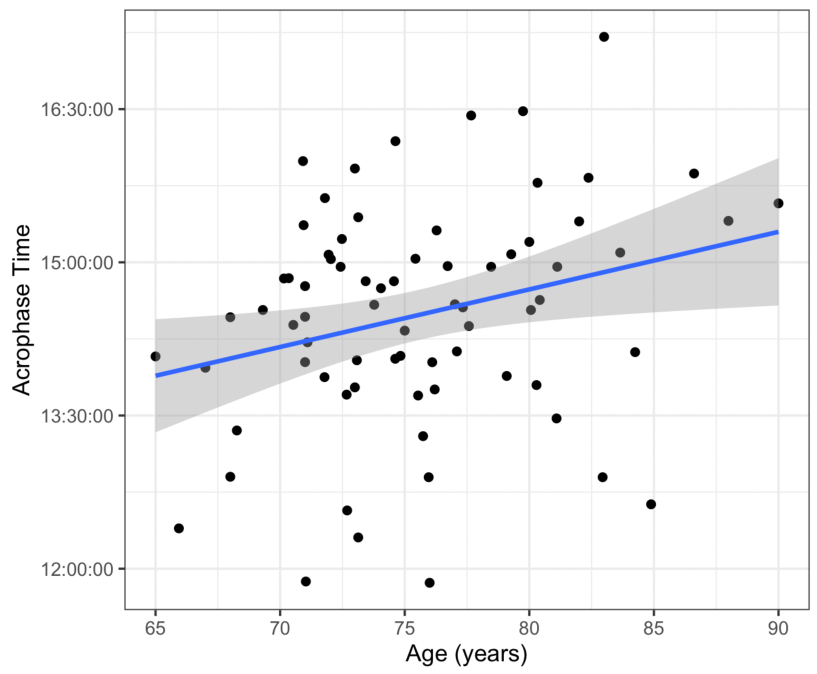
Acrophase time shifts significantly as in older participants. The positive linear relationship between age and acrophase time shows that older individuals tend to have delayed acrophase or peak activity occurring later in the day (R^2^=0.108).

#### Gender and Rest Activity Rhythm Variables

When looking at sex differences in nonparametric RAR, we found no significant associations between variables collected using the AMI Motionlogger (n=35). In the individuals who wore the Philips Spectrum Plus (n=38), however, there were marked differences in interdaily variability (IV) and interdaily stability (IS) between males and females. IS values range from 0 to 1 where 0 is lack of rhythm and 1 is a very stable rhythm. Conversely, higher IV values indicate greater fragmentation of rhythm while lower values convey less fragmentation. We found that cognitively healthy older males had a less stable (*t* (36)=-2.256, *p*=.030) and more fragmented circadian rhythm (*t* (36)=3.514, *p*=.001) than cognitively healthy older females (Figure 3). This finding is supported by the gender differences in CTI-FR and LV scores discussed earlier in this paper where older men were found to be more influenced by circadian type than older women (Table 2). These findings together suggest that circadian behaviors may be more conserved in older women and can be measured effectively both by self-report and objective recording.

**Figure 3.**
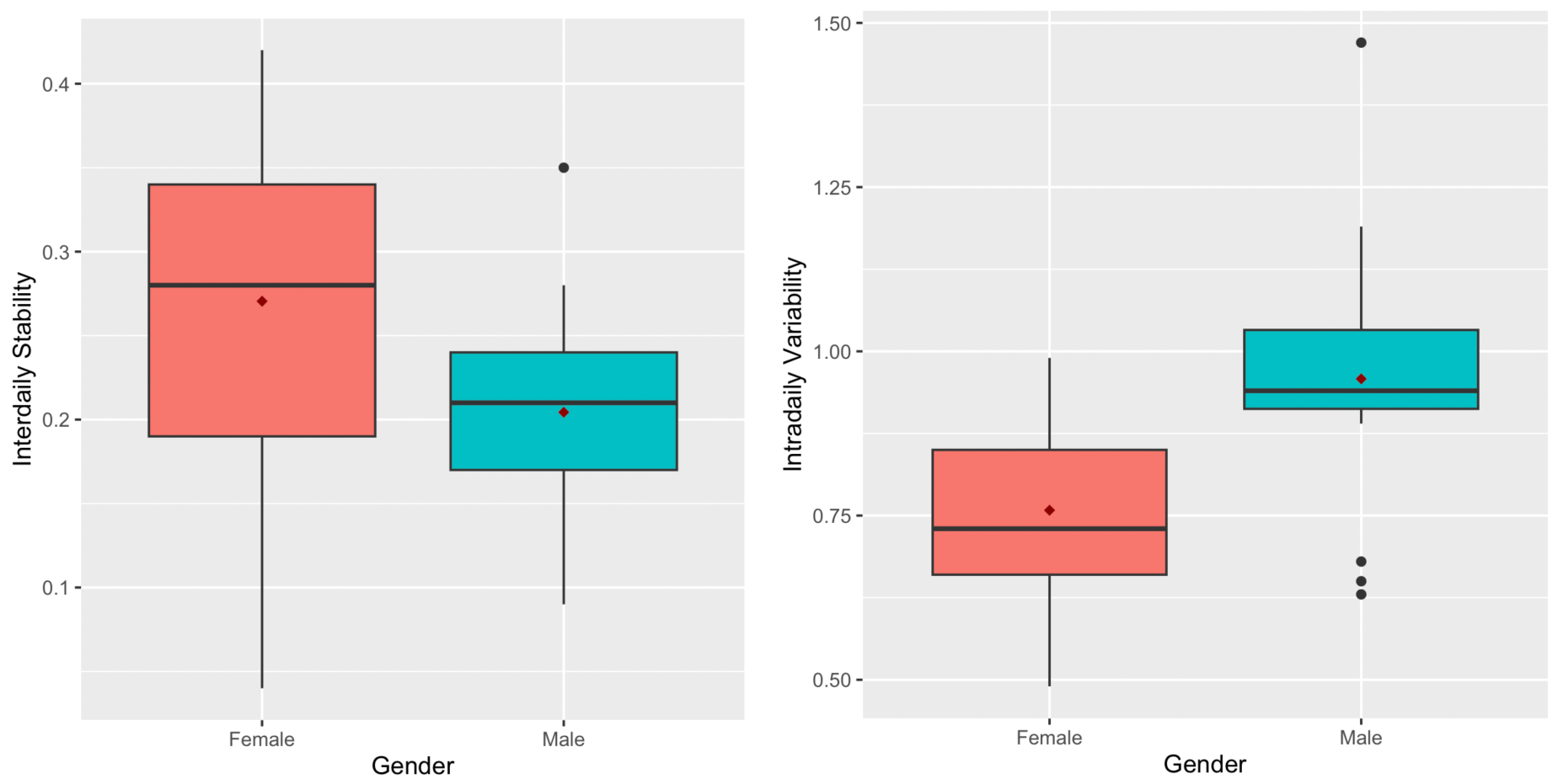
Cognitively healthy older men have lower interdaily stability and higher intradaily variability. In participants wearing the Philips Spectrum Plus Actiwatch, there were significant differences in IS and IV between men and women with older men displaying less stable (MD= −0.0643) and more fragmented (MD= 0.1949) rhythms.

### Relationships Between Self-Reported Circadian Type and Acrophase

Based on the data we collected through actigraphy recordings, we were able to find preliminary evidence for a link between objective and subjective circadian measures. We investigated, specifically, the relationship between nonparametric RAR variables and the CTI and MEQ.

#### CTI and Acrophase

We found a significant association between acrophase time and CTI-FR scores (*F* (4, 62)=3.597, *p*=.011) suggesting that earlier acrophase may be linked to circadian rigidity or resistance to sleep schedule changes (Figure 4). There was a nonsignificant but positive correlation between acrophase time and CTI-LV scores (β=.171, *p*=.147) as well. However, combining the CTI-LV and CTI-FR scores to create the CTI-total score resulted in a stronger correlation between acrophase and these circadian traits, (*F* (4, 62)=2.643, *p*=.042). Earlier acrophase times, indicating an advanced phase, were significantly associated with the combined measures of rigidity and vigor (Figure 5). These findings emphasize that healthy older adults with a peak of activity earlier in the day report more robust circadian behaviors, experiencing ease in overcoming sleepiness and feeling energetic (self-reporting as vigorous people), and they tend to keep a rigid sleep schedule. Conversely, for healthy older adults with a peak of activity that occurs later in the day, it may be harder to overcome sleepiness, rather tending towards lethargy (self-reporting as languid), alongside a susceptibility to variable sleep schedules.

**Figure 4.**
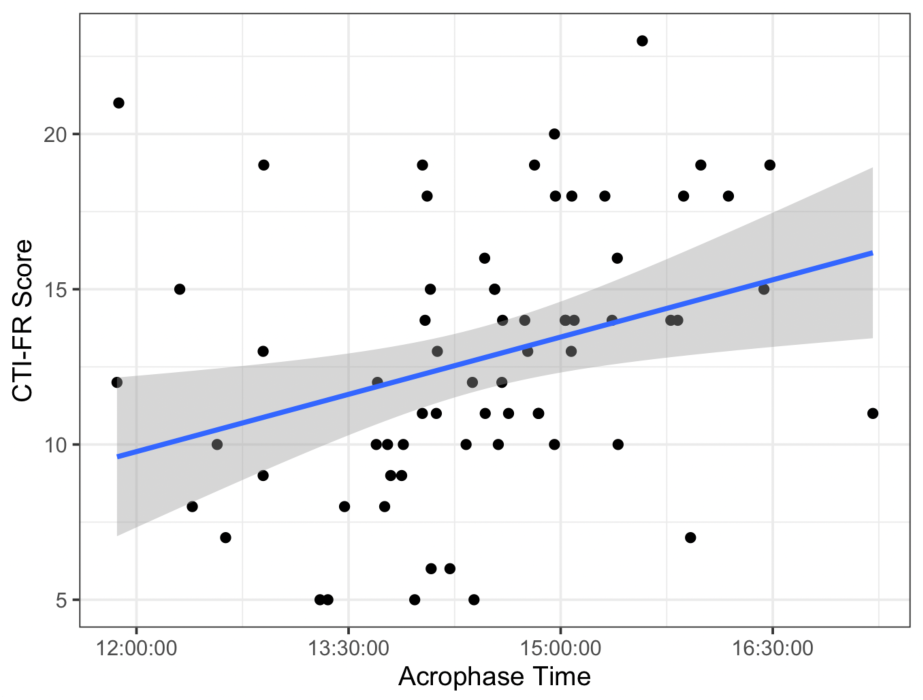
Acrophase associated with CTI-FR scores. Individuals with early acrophase times had lower CTI-FR scores displaying a positive linear relationship between rigid sleep schedules and peak activity earlier in the day (R^2^=0.188, b= .238, p= .052).

**Figure 5.**
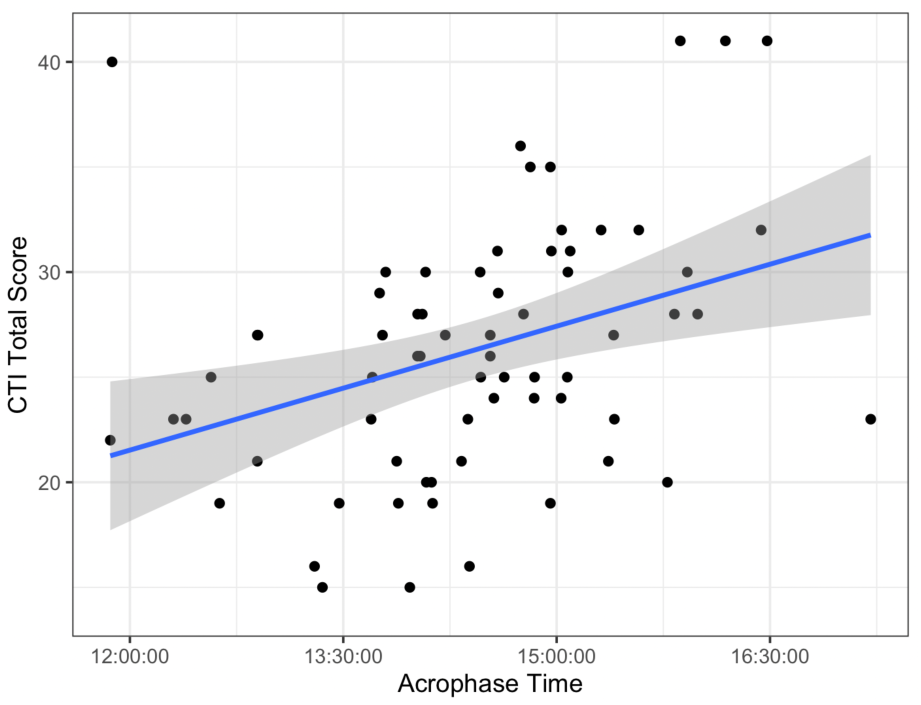
Acrophase time linked to CTI total scores. Older adults self-reporting circadian rigidity and vigor on the CTI have corresponding actigraphy recorded acrophase times supporting early afternoon activity (R^2^=0.146, b= .314, p= .016).

#### MEQ and Acrophase

In our sample of healthy older adults, acrophase time was significantly associated with MEQ scores (*F* (4, 65)=6.535, *p*<.001). Individuals with earlier acrophase times reported a more marked morning preference. In other words, older adults experiencing peak activity earlier in the day tended to be early risers, while older adults with more activity later in the day may be more likely to be night owls (Figure 6).

**Figure 6.**
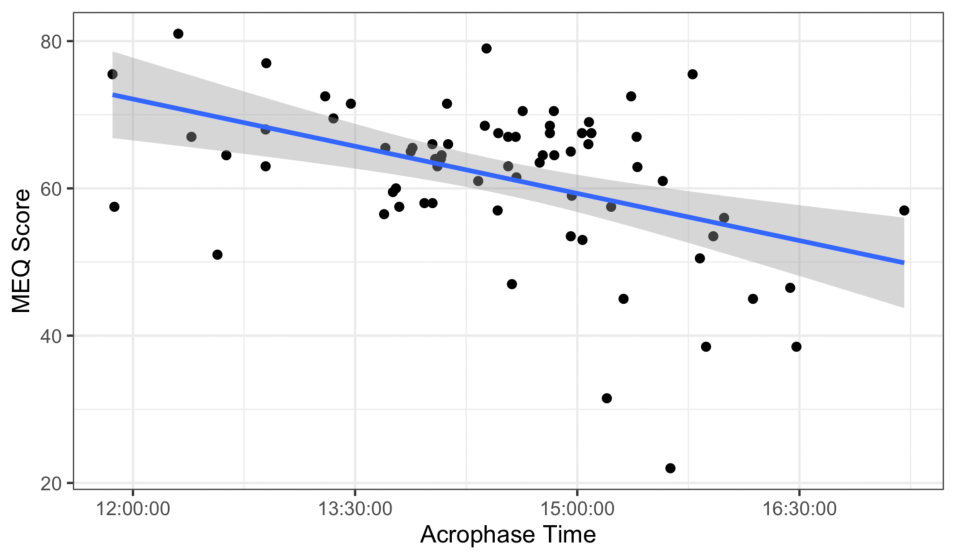
Acrophase time correlated with MEQ scores. Morningness, represented by higher MEQ scores, is associated with early acrophase time suggesting that early risers and those who prefer mornings engage in activity in the day (R^2^=0.287, b= −.442, p<.001).

### Cognitive Function and Morningness-Eveningness

To contextualize the above findings and explore the possible implications of circadian behavior on cognitive processing, we used a multiple linear regression model to examine the relationship between information processing speed (IPS) and the objective and subjective measures discussed above. There were no age or sex associations with IPS, and it was also not linked to CTI-FR and LV scores. We did note a significant association between IPS and morning-evening preference as measured by the MEQ (*F* (4, 51)=3.305, *p*=.018). Faster processing speeds were predictive of higher MEQ scores (β= −.284, *p*=.035) indicating that individuals with faster processing speed tended to be morning people. This finding also implies slower information processing in older adults that prefer evenings and may not be early risers (Figure 7). When looking within sex, IPS differed between men in different time of day preference groups (Table 3) and acrophase times. Moderately Morning Type men, those having a more general preference for mornings, had significantly faster information processing times than Definitely Morning Type, Neither type, or Evening types (η^2^ = 0.25, *p*=.017). Additionally, men with early acrophase times had faster processing speeds than those with delayed phase although this was a weaker correlation (R^2^= 0.453, β =.456, *p*=.073). These findings indicate that morningness in some degree does correlate with improved cognitive processing in healthy older men. There were no significant differences in processing speed across the morning-evening types and no significant correlation between acrophase and cognitive processing in women.

**Figure 7.**
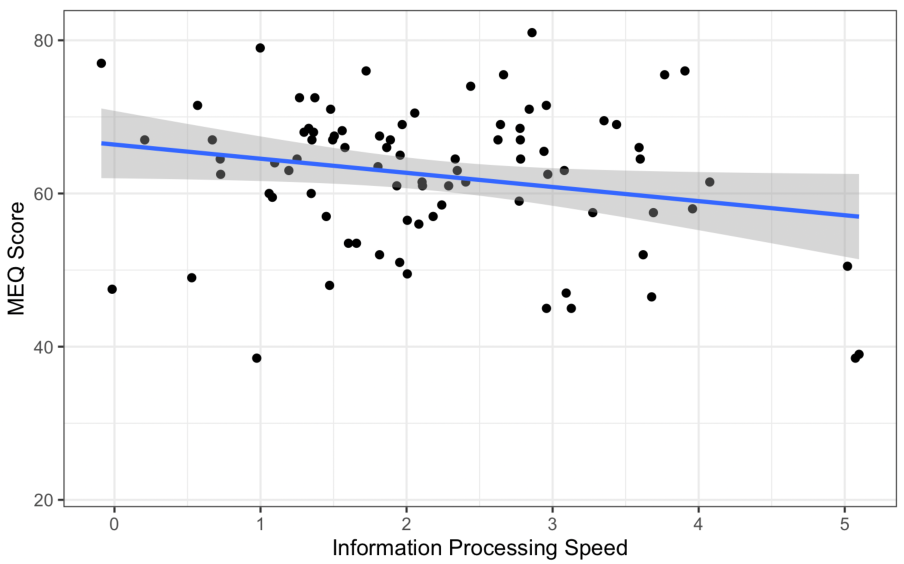
Information processing speed is negatively associated with MEQ scores. Faster response times in the processing speed tasks are linked to higher scores on the MEQ, indicating improved cognitive processing in individuals with morning preferences and self-reported early bird behavior (R^2^=0.211).

## Discussion

The present study reveals several noteworthy patterns, particularly regarding gender differences in circadian behavior and the associations between objectively recorded acrophase and subjective questionnaire responses. The participants in our study were well-educated, cognitively normal older adults with an average age of 75 years and CDR scores of 0 or 0.5. The cohort was skewed towards morning preferences, vigorous circadian types, and rigid sleep schedules, suggesting a propensity for early rising and consistent routines which has been seen in previous studies on older adult populations (Carrier et al., 1997; Marcoen et al., 2015; Monk et al., 1991).

Our findings indicate that sex does impact the relationship between circadian type and cognitive and mood-related variables. Specifically, languid men (high CTI-LV scores) exhibited significantly lower cognitive function, higher depressive symptoms, and a stronger evening preference compared to their vigorous counterparts. Additionally, men with flexible sleep schedules (high CTI-FR scores) had poorer cognitive performance and mood compared to those with rigid schedules. No significant differences were observed between flexible-rigid type or languid-vigorous type women in these domains. This suggests that circadian type may have a more pronounced impact on older men, potentially highlighting greater susceptibility to mood and cognitive impairment linked to circadian misalignment. Gender differences in rest-activity rhythm measures further support the findings in CTI responses. Older men exhibited less stable (low IS) and more fragmented circadian rhythms (high IV) than older women, an association also seen in earlier literature on older adults (Erickson et al., 2024). Gender stratifications in nonparametric RAR variables align with the observed differences in subjective circadian type measures, where men identifying as languid and flexible circadian types were more likely to experience mood and cognitive issues. These results emphasize that circadian behaviors might have distinct implications for cognitive health and mood in older men. However, other studies done only in older women have discovered links between weaker circadian rhythms and worsening cognitive function (Tranah et al., 2011; Walsh et al., 2014). Chronotype changes with menopause have also been assessed using the CSM in women ages 40 to 55, and post-menopausal women were shown to have greater morning preference than pre or peri-menopausal women (Randler & Bausback, 2010). Women with morning preferences also reported more sleep complaints while those with evening preference had more psychological complaints (Randler & Bausback, 2010). This could align with our finding that older men with evening preferences had more mood and cognitive issues than morning-type old men, and our lack of findings in older women could be due to circadian and cognitive changes after menopause that change how older women experience subjective sleep and mood.

The self-reported MEQ scores and objectively measured acrophase times in this cohort align with existing literature suggesting that older adults often exhibit a morning preference as our sample also had a predominantly morning preference and displayed earlier acrophase times (Carrier et al., 1997; Monk et al., 1991). We also observed a correlation between age and morningness suggesting that as older individuals age further their preference might shift from morning to more neutral or evening tendencies. This shift could reflect age-related decomposition of healthy circadian rhythm (Erickson et al., 2024; A. J. Mitchell, 2009). Interestingly, morning preference was associated with faster cognitive processing speeds in men, and moderately morning-type men demonstrated the highest processing speeds. This suggests a potential link between morningness and cognitive efficiency, although this association was not observed in women. The absence of a significant relationship between morning preference and cognitive processing speed in women contrasts with the significant associations found in men, further underscoring the gender differences in circadian behavior and its impact on cognitive function.

Our regression models looking across subjective and objective variables showed that subjective measures of circadian behavior (CTI-LV and CTI-FR) correlated with objective measures of acrophase time. Specifically, individuals with an earlier acrophase time, indicative of a more advanced circadian phase, reported higher vigor and rigidity in their sleep schedules. Delayed acrophase times were associated with more flexible sleep schedules and a tendency towards languidity. Early acrophase times were also positively correlated with morning preferences as measures by the MEQ. These findings emphasize that subjective and objective circadian measures are interrelated, and individuals with an earlier circadian phase tend to exhibit more vigorous behavior, a rigid sleep schedule, and self-identify as “early birds.” This reinforces the importance of circadian alignment in maintaining energetic and structured daily routines. The significant association between faster information processing speeds and morning preference, particularly in men, could suggest better white matter integrity in these individuals, which needs further investigation. Recent literature shows that circadian clock regulation is linked to white matter changes by way of oligodendrocytes and oligodendrocyte precursor cells (OPCs) (Skapetze et al., 2023). Circadian rhythms might play a role in regulating oligodendrocyte function and myelination, and disruptions in circadian patterns could potentially contribute to white matter loss (Colwell & Ghiani, 2019). These findings suggest that slow cognitive processing speeds could be indicative of circadian rhythm disturbances. Thus, the association between faster processing speeds and morning preference in men, highlights the potential cognitive benefits of maintaining a morning-oriented circadian rhythm and suggests that interventions aimed at promoting morningness might enhance cognitive performance in older male adults.

This study, while providing valuable insights into the relationships between gender, circadian rhythms, and subjective and objective measures, is not without limitations. Our sample size was relatively small, predominantly comprised of highly educated, Caucasian individuals which may reduce the generalizability of our findings. Future research should aim to include larger and more diverse cohorts in terms of education, socioeconomic status, and race. The actigraphy data collection relied on two different wrist-based devices, which may have caused variability in the measurements due to differences in device sensitivity and algorithms. While we attempted to account for these differences by including device type as a covariate in our analyses, use of a single, standardized actigraphy device would be a more ideal method. The addition of salivary melatonin assays or core body temperature monitoring to the actigraphy recording would also provide a more accurate and comprehensive assessment of circadian rhythms. Our findings indicated significant sex-related differences in circadian behavior and cognitive function, but the underlying mechanisms driving these differences remain unclear. Future research should focus on exploring the hormonal influences, lifestyle factors, and genetic predispositions which could modulate the sex differences identified, beyond that of sex-at-birth itself. Lastly, the inclusion of neuroimaging, such as DTI, fMRI, or tauPET scans, could provide more insight into which brain regions might be affected by circadian behaviors and whether morning or evening preferences and acrophase times in older adults are linked to neuronal loss and changes in white matter density.

## Data Availability

All data produced in the present study are available upon request and approval from the UCSF Memory and Aging Center.

